# Modifiable lifestyle factors and risk of stroke: a Mendelian randomization analysis

**DOI:** 10.1101/2020.03.17.20037549

**Authors:** Eric L Harshfield, Marios K Georgakis, Rainer Malik, Martin Dichgans, Hugh S Markus

**Affiliations:** Stroke Research Group, Department of Clinical Neurosciences, University of Cambridge, Cambridge, UK; Institute for Stroke and Dementia Research, University Hospital, Ludwig-Maximilians-University, Munich, Germany; Graduate School for Systemic Neurosciences, Ludwig-Maximilians-University, Munich, Germany; Munich Cluster for Systems Neurology (SyNergy), Munich, Germany; German Centre for Neurodegenerative Diseases (DZNE), Munich, Germany

**Author notes:** **Correspondence to:** Dr Eric Harshfield, Department of Clinical Neurosciences, University of Cambridge, R3, Box 83, Cambridge Biomedical Campus, Cambridge, CB2 0QQ, United Kingdom. These authors contributed equally to this work (Eric L Harshfield, Marios K Georgakis). These authors jointly supervised this work (Martin Dichgans, Hugh S Markus).

**Keywords:** Mendelian randomization, education, smoking, body mass index, stroke, genetics

## Abstract

**Aims:** Assessing whether modifiable risk factors are causally associated with reduced stroke risk is important in planning public health measures, but determining causality can be difficult in epidemiological data. Leveraging large-scale genetic data in a technique known as Mendelian randomisation, we aimed to determine whether modifiable lifestyle factors including educational attainment, smoking, and body mass index are causal risk factors for ischaemic stroke and its different subtypes and haemorrhagic stroke.

**Methods and Results:** We performed two-sample and multivariable Mendelian randomization to assess the causal effect of twelve lifestyle factors on risk of stroke and whether these effects are independent. We found genetic predisposition to increased number of years of education to be inversely associated with ischaemic, large-artery, and small-vessel stroke, as well as with intracerebral haemorrhage. Genetic predisposition to ever smoking regularly, higher body mass index (BMI), and higher waist-hip ratio are also associated with ischaemic and large-artery stroke. Additionally, we found that the effects of education, BMI, and smoking on ischaemic stroke to be independent of each other.

**Conclusion:** Genetic predisposition to higher educational attainment can reduce the risk of ischaemic, large-artery, and small-vessel stroke, while genetic predisposition to smoking and higher anthropometry measures can increase the risk of these stroke subtypes. This suggests that lifestyle modification addressing these risk factors will reduce stroke risk.

## INTRODUCTION

Reducing the burden of stroke in the population requires identification of modifiable risk factors, and the demonstration that reducing them reduces stroke risk.^1^ A large number of lifestyle factors have been associated with stroke risk,^2^ but demonstrating whether these associations are causal, and therefore whether modification of a particular risk factor will reduce stroke risk, is less clear. For some environmental factors such as smoking, convincing evidence suggests causality, but for many others it is uncertain whether they are indeed causal. Demonstrating causality is impossible from cross-sectional studies, and can even be challenging in longitudinal epidemiological studies in which risk factor exposure may be captured only over a few years rather than the entire lifespan. One way to assess causality is with a technique known as Mendelian randomization (MR), which uses genetic variants as instrumental variables in an approach analogous to a randomised controlled trial in which allocation to the “treatment group” is determined at conception, to assess whether risk factors have a causal association with an outcome of interest.^3^

A further complicating factor is that stroke represents a syndrome rather than a specific disease, and can be caused by a variety of different pathologies.^4^ The majority (approximately 80%) of strokes are ischaemic, while about 20% are haemorrhagic.^4^ Ischaemic stroke can itself be further divided into aetiological subtypes, of which the main three are cardioembolic (CES), large artery atherosclerotic (LAS), and lacunar or small vessel disease (SVS).^4^ Recent genetic studies indicate that these have distinct genetic and pathophysiological characteristics.^5^ Most epidemiological data examining risk factor modification does not facilitate determination of the effect of modifiable risk factors on specific stroke subtypes.

In this study, we used an MR approach to investigate the aetiological role of modifiable lifestyle risk factors for stroke and its subtypes. We studied a range of modifiable risk factors, including educational attainment, sleep duration, physical activity, smoking status, alcohol consumption, coffee consumption, dietary components, body mass index (BMI), and waist-hip ratio (WHR). As well as examining associations with ischaemic and haemorrhagic stroke, we further investigated the association of these traits with specific ischaemic stroke subtypes (CES, LAS, and SVS). We performed two-sample MR, using genetic variants associated with lifestyle traits as instrumental variables, to determine whether these traits are causally implicated in the risk of stroke. We also performed mediation analysis to evaluate whether the effects of education on ischaemic stroke subtypes are independent to those of BMI and smoking.

## METHODS

### Study design

We performed a two-sample MR analysis, following guidelines for performing and reporting MR studies,^6,7^ using summary statistics from the largest publicly available genome-wide association studies (GWAS) on the following twelve lifestyle factors: educational attainment,^8^ sleep duration,^9^ physical activity,^10^ smoking status,^11^ alcohol consumption,^12^ coffee consumption,^13^ four dietary components,^14^ BMI,^15^ and WHR.^16^ The sample sizes of the included studies ranged from 235,391 to 1.1 million individuals, all of whom were of European ancestry, from cohorts that included UK Biobank, the GIANT consortium, deCODE, and 23andMe. A description of each trait is listed in **Supplementary Table 1**, along with details of the corresponding studies and genetic instruments used.

### Data sources

#### Outcome data

We obtained summary statistics from GWAS of all stroke (AS) and ischaemic stroke subtypes from the MEGASTROKE Consortium,^5^ which consisted of data from 67,162 cases and 454,450 controls. MEGASTROKE had 60,341 cases with any ischaemic stroke (AIS) regardless of subtype, of which 9,006 were cardioembolic stroke (CES), 6,688 were large-artery stroke (LAS), and 11,710 were small vessel stroke (SVS). Stroke cases were defined based on World Health Organization criteria (i.e. sudden onset neurological changes of presumed vascular origin lasting at least 24 hours) with stroke subtypes classified according to the Trial of Org 10172 in Acute Stroke Treatment (TOAST) criteria.^17^ To avoid bias due to population stratification, we restricted our analysis to Europeans since all of the GWAS of the lifestyle traits were also conducted in individuals of European ancestry. From the International Stroke Genetics Consortium (ISGC), we also obtained GWAS summary statistics on intracerebral haemorrhage (ICH),^18^ which consisted of 1,545 cases and 1,481 controls.

#### Instrumental variable selection

For each lifestyle factor included in this analysis, we selected genetic instruments significant at *P* < 1 × 10^−6^ based on the largest published GWAS for that trait with publicly available summary statistics. We also conducted a sensitivity analysis in which we only included single nucleotide polymorphisms (SNPs) that reached genome-wide significance (*P* < 5 × 10^−8^) for association with each risk factor. We then performed linkage disequilibrium (LD) clumping, which ensured that the instruments used for each trait were independent, by selecting only the SNP with the lowest *P*-value amongst all SNPs with an LD *r*^2^ ≥ 0.001.

We obtained data on educational attainment from a GWAS of time spent in education measured in years, from which we selected 440 SNPs. The study included 1.1 million individuals from 71 cohorts (including UK Biobank, 23andMe, and deCODE).^8^ The GWAS of sleep duration analysed sleep duration measured in hours in 446,118 individuals from UK Biobank, adjusted for age, sex, 10 principal components of ancestry, genotyping array, and genetic correlation matrix, with 125 SNPs selected as genetic instruments.^9^ The GWAS of physical activity analysed moderate-to-vigorous physical activity, which was measured in MET-minutes per week with adjustment for age, sex, genotyping chip, first 10 genomic principal components, centre, and season, in 377,234 individuals from UK Biobank, and we selected 7 SNPs as instruments from this study.^10^ The GWAS of smoking status used a lifetime smoking index, which was constructed in 462,690 individuals from UK Biobank by developing a model that incorporated self-reported time started smoking, duration of smoking, and cigarettes per day, as well as half-life and lag time constants to capture the non-linear risk of smoking on health.

We selected 210 SNPs as instruments for the lifetime smoking index.^11^ The GWAS of alcohol consumption was based on the number of drinks consumed per week in 941,280 individuals from a number of cohorts including UK Biobank, 23andMe, and deCODE, from which we selected 83 SNPs.^12^ The GWAS of coffee consumption was based on the number of cups of coffee consumed per day as reported by 336,448 UK Biobank participants on a touchscreen questionnaire at the assessment centre, with adjustment for age, sex, BMI, total energy, proportion of 24-hour recalls self-reported as capturing “typical intake,” and top 20 principal components, for which our genetic instrument had 49 SNPs.^13^ The GWAS of dietary components consisted of four traits (fat, protein, carbohydrate, and sugar) measured in up to 268,922 participants from UK Biobank, the DietGen consortium, and several other cohorts, with 5, 7, 8, and 9 SNPs respectively as genetic instruments.^14^ The GWAS of BMI was performed in 681,275 individuals from UK Biobank and the Health and Retirement Study, with adjustment for age, sex, recruitment centre, genotyping batches, and 10 principal components,^15^ and the GWAS of WHR was performed in 694,649 individuals from UK Biobank and the GIANT consortium with adjustment for SNP array.^16^ These studies included 623 and 380 SNPs respectively as instruments.

### Statistical analyses

We used the inverse-variance weighted (IVW) method as the primary MR analyses, which uses inverse-variance-weighted meta-analysis under a random-effects model to combine the ratio estimates from each genetic variant into a single estimate of the causal effect of the lifestyle trait on the outcome.^3^ We also conducted sensitivity analyses using a variety of robust MR methods, which employ different sets of assumptions to make reliable causal inferences. These approaches included MR-Egger regression, the weighted median estimator, and the simple and weighted mode-based estimators.^3^

For each trait, we harmonised all SNPs associated with the trait with the outcome data to ensure that the effect estimates of each SNP on the exposure and on the outcome corresponded to the same effect allele. We then performed MR using the IVW method and sensitivity analyses using additional MR approaches. We accounted for multiple testing using a false discovery rate (FDR) cut-off of *q* < 0.05. Analyses were performed in R version 3.6.1 (R Core Team, 2019) using the TwoSampleMR package version 0.4.25.^19^ Two-sided *P*-values and 95% confidence intervals are presented.

We also performed MR-based network mediation analysis^20^ to examine the extent to which increases in BMI and increased levels of smoking mediate the protective effective of education on ischaemic stroke subtypes. We calculated the total effect of education on stroke using two-sample univariable random-effects MR using the IVW approach. To calculate the direct effects, we combined the instruments for education, BMI, and smoking, and performed multivariable MR analyses to estimate their associations with the stroke subtypes. We calculated the indirect effect of education on stroke that acts through either BMI or smoking as the difference between the direct and total effect estimates. Finally, we calculated the proportion of the total effect of education on stroke that was mediated by BMI and smoking by dividing the indirect effect by the total effect. We also used the delta method^21^ to derive standard errors for each of the effects.

## RESULTS

We analysed twelve lifestyle traits for association with AS, AIS, ICH, and the three subtypes of ischaemic stroke (CES, LAS, and SVS). **Figure 1** summarises the direction and magnitude of the association estimates for each lifestyle trait with each outcome, with stronger associations indicated by darker colours (red in the positive direction and blue in the negative direction) and asterisks to indicate the level of statistical significance. **Figure 2** shows scatterplots of the associations of each genetic variant plotted against their association with corresponding outcomes for all traits that had significant (FDR *q* < 0.05) associations. Forest plots of associations between lifestyle traits and each stroke subtype using the random-effects inverse-variance weighted method are shown in **Figure 3**. The full results for the association of each of the twelve traits with all six outcomes, using each MR method, are provided in **Supplementary Table 2**.

**Figure 1.**
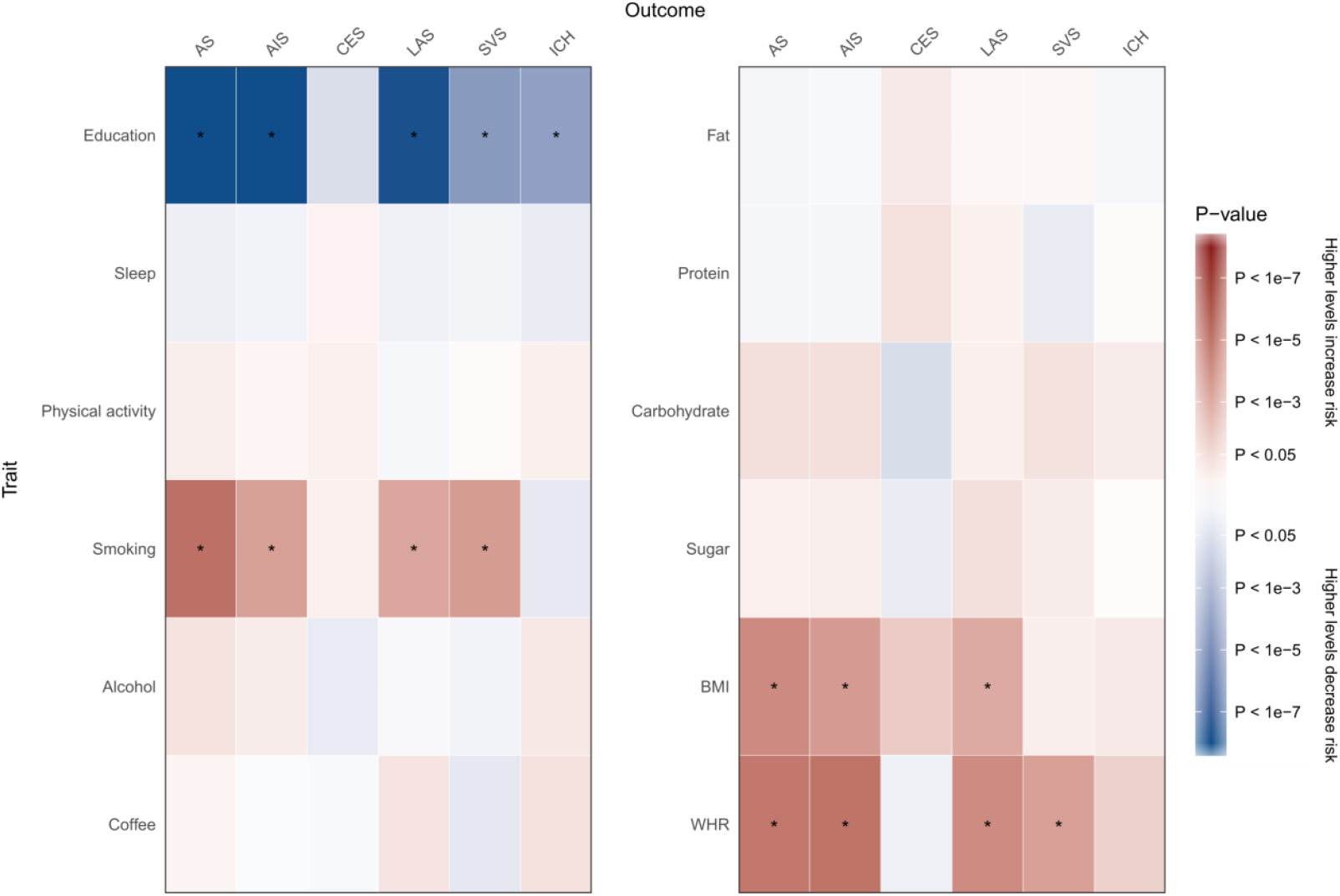
Mendelian randomization results showing causal estimates for association of lifestyle traits with stroke and its subtypes. Colours show magnitude and direction of *P*-value of association for estimate of causal effect using inverse-variance weighted MR approach. *P*-values less than 1 × 10^−8^ have been truncated in the figure. Asterisks indicate significant associations for the causal effect estimates (FDR *q* < 0.05). **AS** = All stroke; **AIS** = Any ischaemic stroke; **CES** = Cardioembolic stroke; **ICH** = Intracerebral haemorrhage; **LAS** = Large-artery stroke; **SVS** = Small vessel stroke. Please refer to **Supplementary Table 1** for a description of each trait.

**Figure 2.**
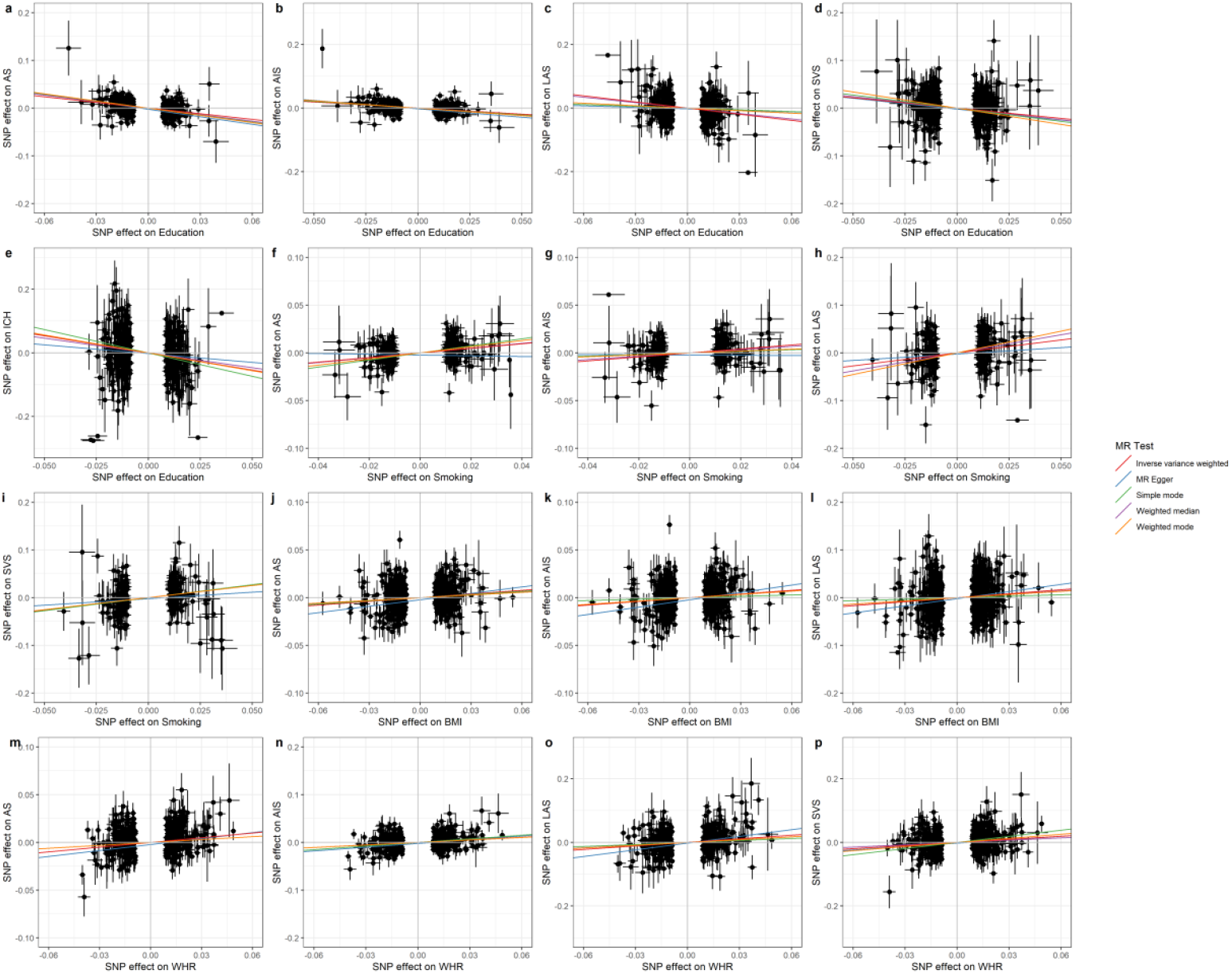
Genetic associations of lifestyle traits and stroke subtypes for significant causal estimates. The associations of each genetic variant associated with lifestyle traits with significant (FDR *q* < 0.05) causal estimates are plotted against their association with the corresponding outcome. Circles represent the associated change in levels of the trait and corresponding increased risk for each variant. The horizontal and vertical lines through each circle represent the corresponding 95% confidence intervals for the genetic associations. Associations were oriented to the effect allele of each trait. Coloured lines show the slope (causal estimate) of the trait on the outcome obtained using a variety of different MR approaches. Trait / Outcome: (a) Education / AS; (b) Education / AIS; (c) Education / LAS; (d) Education / SVS; (e) Education / ICH; (f) Smoking / AS; (g) Smoking / AIS; (h) Smoking / LAS; (i) Smoking / SVS; (j) BMI / AS; (k) BMI / AIS; (l) BMI / LAS; (m) WHR / AS; (n) WHR / AIS; (o) WHR / LAS; (p) WHR / SVS. Please refer to **Supplementary Table 1** for a description of each trait.

**Figure 3.**
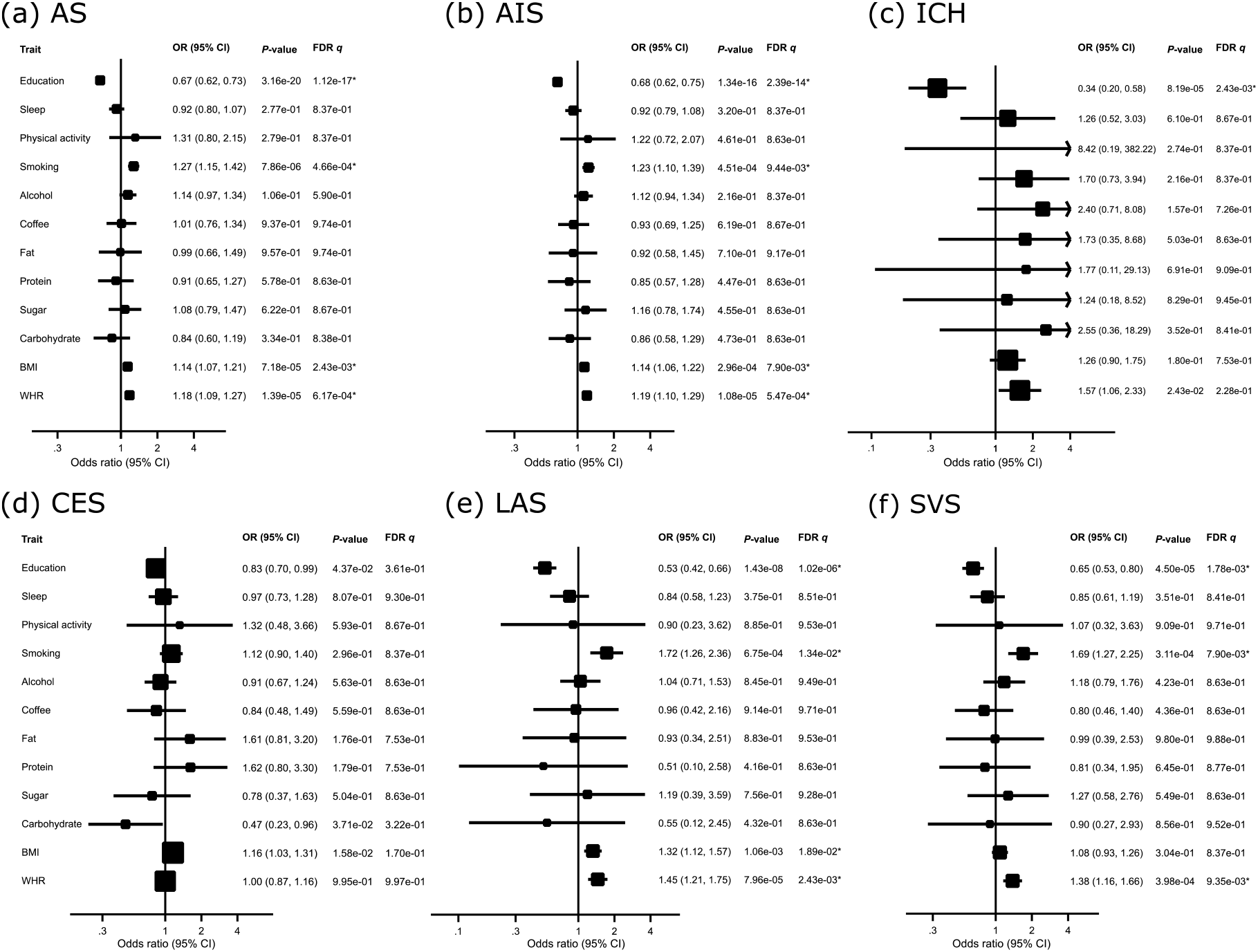
Mendelian randomization associations between genetic predisposition to lifestyle factors and stroke subtypes. Results derived from random-effects inverse-variance weighted MR analyses. Asterisks indicate significant associations for the causal effect estimates (FDR *q* < 0.05). Results are shown for (a) all stroke, (b) any ischaemic stroke, (c) intracerebral haemorrhage, and ischaemic stroke subtypes: (d) cardioembolic stroke, (e) large-artery stroke, and (f) small vessel stroke. Please refer to **Supplementary Table 1** for a description of each trait.

We found significant inverse associations of genetically determined number of years of education with ischaemic stroke (OR: 0.68, 95% CI: 0.63-0.75) and intracerebral haemorrhage (OR: 0.34, 0.20-0.58), as well as with large-artery (OR: 0.53, 0.42-0.67) and small-vessel stroke (OR: 0.65, 0.53-0.80). There was also suggestive (but not statistically significant) evidence of an inverse association of education with cardioembolic stroke (OR: 0.83, 0.70-0.99).

Genetic predisposition to lifetime smoking was significantly associated with ischaemic stroke (OR: 1.23, 1.10-1.39), large-artery (OR: 1.72, 1.26-2.36) and small-vessel stroke (OR: 1.69, 1.27-2.25), but not with cardioembolic stroke (OR: 0.83, 0.70-0.99). The association with intracerebral haemorrhage was of similar magnitude but did not reach statistical significance (OR: 1.70, 0.73-3.94).

Genetically determined higher BMI was significantly associated with increased risk of ischaemic (OR: 1.14, 1.06-1.22) and large-artery stroke (OR: 1.32, 1.12-1.57), and there was a suggestive association with cardioembolic stroke but it was not statistically significant (OR: 1.16, 1.03-1.31). Genetically higher WHR was significantly associated with increased risk of ischaemic stroke (OR: 1.19, 1.10-1.29), large-artery (OR: 1.45, 1.21-1.75) and small-vessel stroke (OR: 1.38, 1.16-1.66), but showed no association with cardioembolic stroke (OR: 1.00, 0.87-1.16). There were no material differences in the results when the instruments were restricted to SNPs that reached genome-wide significance for association with each trait (**Supplementary Table 3**).

There were no significant associations between sleep duration, physical activity, dietary components, coffee intake, or alcohol consumption with ischaemic stroke, intracerebral haemorrhage, or any ischaemic stroke subtype.

We calculated that the proportion of the total effect of education on stroke mediated through BMI and smoking was 1% for ischaemic stroke and nearly 14% for cardioembolic and large-artery stroke. However, the indirect effects of education on each of the five stroke subtypes were not statistically significant (*q* < 0.05), and for all stroke and small-vessel stroke there was inconsistent mediation because the indirect effects and total effects were in opposing directions, which may be due to insufficient power. Our mediation analysis therefore lacked sufficient evidence to state that the effects of education on ischaemic stroke and its subtypes are mediated through either BMI or smoking. This suggests that education, BMI, and smoking each have independent effects on stroke.

## DISCUSSION

In this study, we conducted MR analyses using large-scale GWAS summary statistics to investigate causal associations of modifiable lifestyle traits with risk of stroke subtypes. We found genetic predisposition to number of years of education to be inversely associated with ischaemic stroke, large-artery and small-vessel stroke, and intracerebral haemorrhage. Genetic predisposition to ever smoking regularly and having higher BMI and WHR were associated with ischaemic and large-artery stroke. We also found that the associations of education, BMI, and smoking on risk of ischaemic stroke and its subtypes were independent of each other.

Out analysis did not identify any evidence of a causal association of lifestyle factors with cardioembolic stroke. This may reflect the fact that the majority of cases of CES are due to atrial fibrillation,^22^ which has a different genetic and other risk factor profile to other stroke subtypes. While further research into the underlying mechanisms is needed, this could help explain why we found that lifestyle factors are implicated in the onset of large-artery and small-vessel stroke but not CES.

Our MR results provide genetic evidence for an inverse causal effect of educational attainment on ischaemic stroke and intracerebral haemorrhage. Our analysis validates previously reported associations of the effect of education on ischaemic stroke from observational studies^23^ and MR analyses,^24^ as well as a recent MR analysis showing associations of education with large-artery stroke, small-vessel stroke, and intracerebral haemorrhage.^25^ Our findings for the association of the lifetime smoking index with increased risk of ischaemic, large-artery, and small-vessel stroke is in concordance with previous observational studies^26^ and MR analyses.^27^ While earlier MR analyses of BMI with smaller sample sizes did not show clear evidence of a causal association with any ischaemic stroke subtype,^28^ our analysis in a larger sample confirms the associations reported in a recent MR analysis of BMI with large-artery stroke and WHR with large-artery and small-vessel stroke.^29^

We did not identify any associations of stroke or its subtypes with genetic predisposition to longer sleep duration, increased physical activity, or increased consumption of fat, protein, carbohydrates, sugar, coffee, or alcohol. The number of genetic instruments was very small for physical activity and the dietary components (fewer than 10 SNPs each), which may have resulted in weak instrument bias. Additionally, analysing each dietary component in isolation may be less effective than considering the total effect of a healthy or unhealthy diet on stroke and its subtypes. Nevertheless, our research shows that sleep, physical activity, and several diet-related factors do not exhibit evidence of a causal association with ischaemic stroke and its subtypes or intracerebral haemorrhage. In contrast, genetic liability to insomnia has been shown to be associated with ischaemic stroke in a recent MR analysis,^30^ so it is possible that overall sleep duration is not specific enough as a phenotype.

Our findings lend support to a number of potential policy recommendations. While some individuals may have a genetic predisposition to lower educational attainment, higher frequency of smoking, or higher BMI or WHR, genetic heritability only explains a fraction of the total variation in these traits (11% for education, 2% for smoking, 6% for BMI, and 3.9% for WHR).^8,11,15,16^ Therefore, regardless of their level of genetic risk, individuals can make various lifestyle changes to reduce their risk of stroke, such as smoking cessation.^2,31,32^ This study also highlights the importance of promoting universal education, which would benefit everyone regardless of genetic makeup.^25^ Interestingly, although BMI and WHR were significantly associated with increased risk of stroke, we did not identify significant associations for related lifestyle changes that could be adopted to help reduce adiposity measures, such as increased physical activity and reduced fat and sugar consumption. This may be due to the small number of genetic variants that were available to use as instrumental variables for physical activity and dietary components.

Our study has several strengths. The large sample size of each study (ranging from 235,000 to 1.1 million participants) provided substantial power to detect differences in the effects of lifestyle factors on aetiological stroke subtypes and to perform multiple sensitivity analyses to test the validity of the MR assumptions, which reduced the likelihood of obtaining biased estimates. In addition, our investigation of a wide range of lifestyle factors provides a relatively comprehensive overview of the associations with subtypes of ischaemic stroke and intracerebral haemorrhage. Moreover, several of the pitfalls common to observational studies, such as reverse causation bias and potential confounding, were reduced or avoided altogether by using an MR approach since genetic variants are allocated at conception. Our estimates therefore represent the impact of a lifelong intervention in the lifestyle factors rather than lifestyle changes of a shorter duration as reported in observational studies.

Our study also has several limitations. Under the MR analysis framework, we used genetic variants as instrumental variables to evaluate whether modifiable lifestyle factors have a causal association with risk of stroke and its subtypes; however, as we only had access to summary-level data rather than individual participant data, we were not able to calculate polygenic risk scores to determine whether individuals with specific genetic variants were at increased risk of stroke. A related point is that while we did capture the impact of a lifelong intervention in each lifestyle factor, our analyses only considered the genetic predisposition to adopt a certain lifestyle habit rather than determining whether individuals who actually implement a given change to their lifestyle have a measurable impact on their risk of stroke, which may also be influenced by various environmental factors. Additionally, our analyses were based on datasets involving individuals of European ancestry and thus might not be applicable to other ethnicities. Furthermore, although the association of BMI with stroke is J-shaped,^33^ our analyses did not account for non-linear associations for continuous traits.

In conclusion, our results suggest causal associations of lower educational attainment and higher levels of smoking, BMI, and WHR with increased risk of ischaemic stroke, particularly large-artery and small-vessel stroke. The effects of modifiable lifestyle factors on large-artery and small-vessel stroke observed here may have important policy implications that support the implementation of strategies to encourage improved lifestyle habits.

## Data Availability

The GWAS summary statistics used to perform the analyses described in this study were obtained from publicly available published data. All data generated or analysed during this study are included in the manuscript and its supplementary materials.

https://www.thessgac.org/data

http://sleepdisordergenetics.org/informational/data

https://data.bris.ac.uk/data/dataset/10i96zb8gm0j81yz0q6ztei23d

https://conservancy.umn.edu/handle/11299/201564

https://digitalhub.northwestern.edu/collections/7badb7c9-d4ec-4ca9-b58e-6e01f224fcf7

https://portals.broadinstitute.org/collaboration/giant/index.php/GIANT_consortium_data_files

https://www.megastroke.org/

http://cerebrovascularportal.org/

## FUNDING

This work was supported by the British Heart Foundation [RG/16/4/32218]; the European Union’s Horizon 2020 research and innovation programme [No 667375 (CoSTREAM) and No 666881 (SVDs@target)]; a grant for strategic collaboration between Ludwig-Maximilians-University Munich and the University of Cambridge; Cambridge University Hospitals National Institute for Health Research Biomedical Research Centre [*]; the Onassis Foundation [to M.K.G.]; the Deutsche Forschungsgemeinschaft [Munich Cluster for Systems Neurology (EXC 1010 SyNergy) and Collaborative Research Center 1123 (B3) to M.D.]; the Corona Foundation [to M.D.]; Fondation Leducq [Transatlantic Networks of Excellence Program on the Pathogenesis of Small Vessel Disease of the Brain to M.D.]; and the National Institute for Health Research [Senior Investigator Award to H.S.M.] [*]. The funding organisations had no role in any of the following: design and conduct of the study; collection, management, analysis, and interpretation of the data; review, or approval of the manuscript. The corresponding author had full access to all the data in the study and had final responsibility for the decision to submit for publication. [*] The views expressed are those of the authors and not necessarily those of the NHS, the NIHR, or the Department of Health and Social Care.

## ACKNOWLEDGEMENTS

This research was conducted using the UK Biobank resource under application number 36509. We acknowledge the contributions of the MEGASTROKE consortium, the International Stroke Genetics Consortium (ISGC) Cerebrovascular Disease Knowledge Portal, the Social Science Genetic Association Consortium (SSGAC), the Sleep Disorder Knowledge Portal (SDKP), and the Genetic Investigation of ANthropometric Traits (GIANT) consortium for making their datasets publicly available. The MEGASTROKE project received funding from sources specified at http://megastroke.org/acknowledgements.html, and the full investigator list of the MEGASTROKE consortium is available at http://megastroke.org/authors.html.

## Author contributions

E.L.H., M.K.G., H.S.M., and M.D. conceived and designed the study. E.L.H. and M.K.G. analysed the data. All authors participated in interpretation of the results. E.L.H. and H.S.M. drafted the manuscript. All authors critically revised the manuscript for intellectual content and approved the final submitted version.

## Conflicts of interests

None reported.

## Notes

### Competing Interest Statement

The authors have declared no competing interest.

